# A systematic review of medical and clinical research landscapes and quality in Malaysia and Indonesia [REALQUAMI]: the review protocol

**DOI:** 10.1101/19004010

**Authors:** Boon-How Chew, Lim Poh Ying, Shaun Wen Huey Lee, Navin Kumar Devaraj, Adibah Hanim Ismail @ Daud, Nurainul Hana Shamsuddin, Puteri Shanaz Jahn Kassim, Aneesa Abdul Rashid, Aaron Fernandez, Noraina Muhamad Zakuan, Soo Huat Teoh, Akiza Roswati Abdullah, Hanifatiyah Ali, Abdul Hadi Abdul Manap, Fadzilah Mohamad, Indah S. Widyahening

**Author notes:** Correspondence: Boon-How Chew, Department of Family Medicine, Faculty of Medicine & Health Sciences, Universiti Putra Malaysia, 43400 Serdang, Selangor, Malaysia.

## Abstract

**Background:** Research landscapes and quality may change in many ways. Much research waste has been increasingly reported. Efforts to improve research performance will need good data on the profiles and performance of past research.

**Purpose:** To describe the characteristics and quality of clinical and biomedical research in Malaysia and Indonesia.

**Methods:** A search will be conducted in PubMed, EMBASE, CINAHL and PsycINFO to identify for published clinical and biomedical research from 1962 to 2017 from Malaysia and/or Indonesia.

Additional search will also be conducted in MyMedR (for Malaysian team only). Studies found will be independently screened by a team of reviewers, relevant information will be extracted and quality of articles will be assessed. As part of quality control, another reviewer will independently assess 10-20% of the articles extracted. In Phase 1, the profiles of the published research will be reported descriptively. In Phase 2, a research quality screening tool will be validated to assess research quality based on three major domains of relevance, credibility of the methods and usefulness of the results. Associations between the research characteristics and quality will be analysed. The independent effect of each of the determinant will be quantified in multivariable regression analysis. Longitudinal trends of the research profiles, health conditions in different settings will be explored. Depending on the availability of resources, this review project may proceed according to the different clinical and biomedical disciplines in sequence.

**Discussion:** Results of this study will serve as the ‘baseline’ data for future evaluation and within country and between countries comparison. This review may also provide informative results to stakeholders of the evolution of research conduct and performance from the past till now. The longitudinal and prospective trends of the research profiles and quality could provide suggestions on improvement initiatives. Additionally, health conditions or areas in different settings, and whether they are over- or under-studied may help future prioritization of research initiatives and resources.

## Introduction

There is now an increasing number of clinical and biomedical research and publications published, especially those originating from Asia. Huge research wasting have been reported because of irrelevancy,^1^ poor research designs,^2^ inaccessible research data ^3^ and incomplete reporting.^4,5^ Moreover, “It was very easy to make errors” as admitted by John Ioannidis, one of the co-director at the new Meta-Research Innovation Center at Stanford (METRICS) on the challenges along the research process despite the noble intentions of the researchers.^6^ However, it is uncertain of the actual clinical and biomedical research landscapes that is evolving throughout the past decades in Asia. Similarly, the quality of the published research in a country such as Malaysia over the past few decades has not been examined. These assessment and evidence are needed to inform the existing researchers, research institutes and funders in Malaysia of adequacy of current effort or a need to improvise the existing ways of conducts.

There are about 200 tools available for evaluating research quality or biases in randomized and non-randomized studies.^7-9^ Nevertheless, most tools available for assessing non-randomized studies are generally of poor methodological quality, making that the assessment of methodological quality and risk of bias consistently across primary studies difficult or impossible.^10^ Many different tools exist for different study designs such as the Cochrane Risk of Bias tool for randomized trials,^11^ the QUADAS 2 tool ^12^ for diagnostic test accuracy studies, and the AMSTAR ^13^ and ROBIS tools ^14^ for systematic reviews, and the ROBINS-I ^15^ for non-randomized studies of the effects of interventions. Additionally, there are a few web-based tools and checklist for different study designs such as the NIH Study Quality Assessment Tool for controlled intervention studies, systematic reviews and meta-analyses, observational cohort and cross-sectional studies, case-control, pre-post, case series studies (https://www.nhlbi.nih.gov/health-topics/study-quality-assessment-tools); the Critical Appraisal Skills Programme (CASP) checklists by an Oxford-based Better Value Healthcare Ltd (https://casp-uk.net/casp-tools-checklists/); a web application Critical Appraisal Tools (FLC 2.0) developed by OSTEBA Spain to guide critical appraisal process (http://www.lecturacritica.com/es/acerca.php).

Among some of the more widely used and recommended tools are the Newcastle-Ottawa scale,^16^ the Downs and Black instrument ^17^ and the latter RTI item bank (RTI-IB).^18^ The Newcastle-Ottawa Scale (NOS), which has been used to illustrate issues in data extraction from primary non-randomized studies, and it has only eight items and is simpler to apply.^16^ However, the items may still need to be customized to the review question of interest. The Downs and Black instrument ^17^ has been modified for use in a methodological systematic review.^8^ The reviewers found that some of the 29 items were difficult to apply to case control studies, that the instrument required considerable epidemiological expertise and that it was time consuming to use. There are reports that these tools are difficult to apply,^19-22^ and agreement between review authors is modest. Median observed inter-rater agreement for the RTI-IB was 75% (25th percentile [p25] =61%; p75 =89%), median first-order agreement coefficient statistic was 0.64 (p25 =0.51; p75 =0.86). Although the RTI-IB facilitates a more complete quality assessment than the NOS but is more burdensome. Additionally, there are different meanings in epidemiological terminology in different countries for example the term ‘selection bias’ describes what others may call ‘applicability’ or ‘generalizability’. Thus, comprehensive manuals are required to accompany these tools to offer instructions for standardized interpretation by different users. However, this may pose a great challenge to users and not many tools have such comprehensive manual.

Therefore, no tool is found adequate as an all-rounded tool for all types of study designs,^9^ or is a recommended tool that is suitable to assess the quality of the published researches as a relatively quick screening tool. Accordingly, we assimilate the quality indicators used in the existing tools, based on the series of the users’ guides to the medical literature by the Evidence-Based Medicine Working Group ^23,24^ and a recent review ^25^ and principles of clinical epidemiology,^26^ and developed one for this review project.

### Aims of the project

This project aims to systematically identify for published research articles performed by researchers in each participating country. For example, we aim to identify for articles published by Malaysian researchers on research conducted in Malaysia. We will subsequently assess the characteristics and quality of the researches published in journals as described below.

## Material and methods

This systematic review will consists of two phases. In the first phase, we will descriptively report the demographics and characteristics of studies performed in each country to date (research landscapes). In the second phase, we will assess the quality of the research based on the published reports in journals (research quality) (Figure 1).

**Figure 1.**
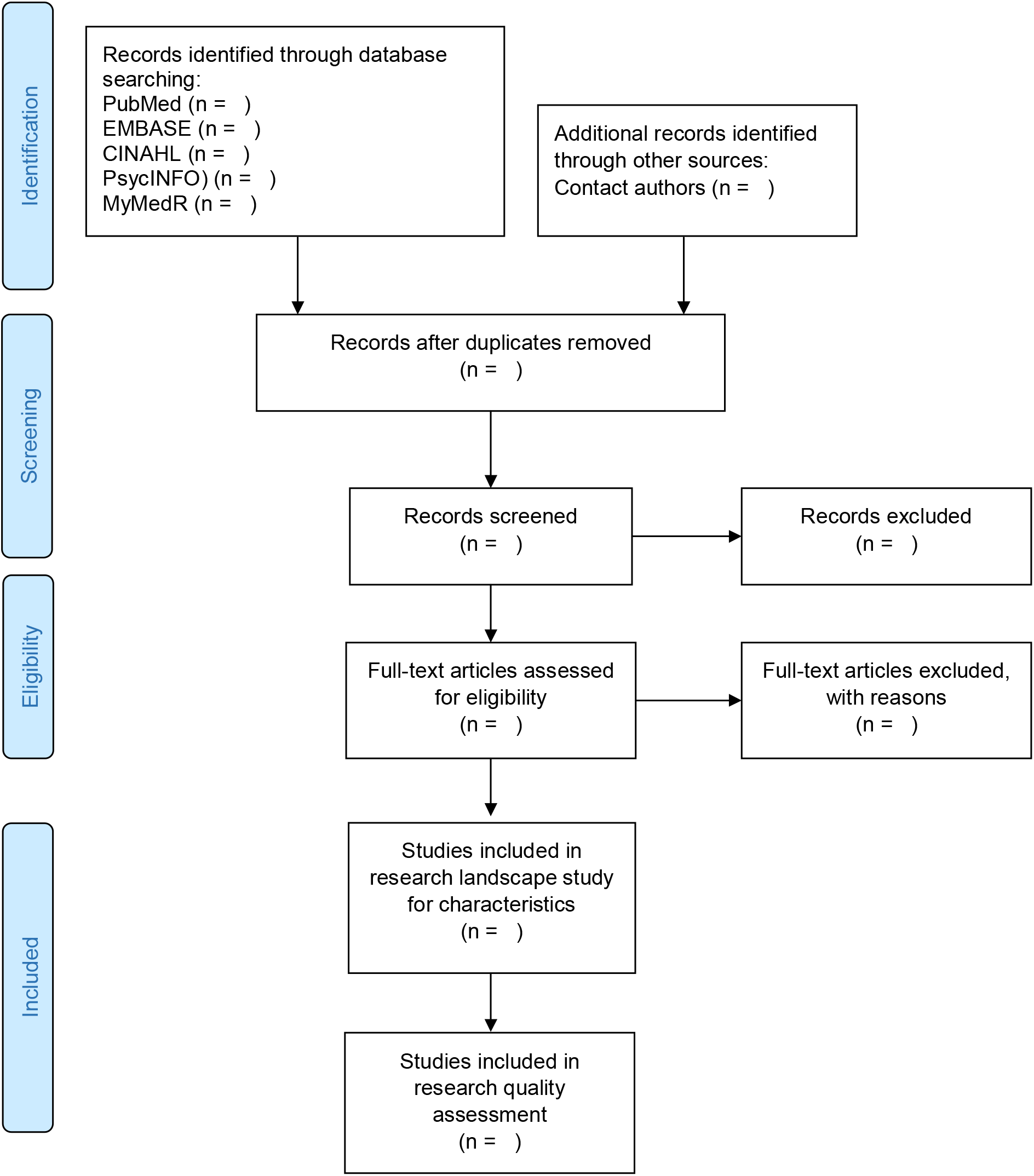
The two phases of the review. Adapted from: Moher D, Liberati A, Tetzlaff J, Altman DG, The PRISMA Group (2009). Preferred Reporting Items for Systematic Reviews and Meta-Analyses: The PRISMA Statement. PLoS Med 6(7): e1000097. doi:10.1371/journal.pmed1000097. For more information, visit www.prisma-statement.org.

### Protocol and registration

A protocol of this systematic review is available from the Open Science Framework’s registry for Research on the Responsible Conduct of Research (RRRCR) with the registration ID: https://osf.io/w85ce.

### Inclusion criteria and search strategy

All clinical and biomedical research conducted in Malaysia or Indonesia from January 1962 (Malaysia after Singapore independence) to December 2017 will be identified from the following databases: PubMed, EMBASE, CINAHL and PsycINFO. We will include all published papers of health and biomedical researches done in each country (Malaysia or Indonesia) or by citizen of each country (Malaysian or Indonesian) with an affiliation in one of the institution in each country (Malaysian or Indonesian). We will also search for additional literature from MyMedR (http://mymedr.afpm.org.my/) database as it specifically compiles published papers in health and biomedical research conducted in Malaysia or by authors who has a Malaysian affiliation as well as from MyJurnal, an online system used by Malaysia Citation Centre (MCC), Ministry of Higher Education Malaysia to collect and index all the Malaysian journals. Search results will be compiled into Endnote reference management software where duplicates will be removed. If necessary, authors and institutions will be contacted. A medical librarian and a science officer at the Faculty of Medicine and Health Sciences Universiti Putra Malaysia will complete these tasks. The review work will be completed by two separate teams with each is based in Malaysia and Indonesia, respectively.

### Study selection and data extraction

All review authors will independently screen identified articles by title and abstract. Full text of eligible article will be retrieved and independently extracted using a standard data extraction template. This template will be pilot-tested on 10 articles among all the review authors for clarity, and modification of the template will be done accordingly. The final piloted template is available as Supplementary Table 1. To ensure the data quality, a review author (BHC) will reassess 10-20% of the articles. Any discrepancy will be solved by consensus between three or more review authors.

In the event of duplicate publications or multiple reports of a research study, we will use the most complete data set aggregated across all known publications. Duplicate publications are defined as two or more published articles that report on the same research question. All review authors will learn about the principles of clinical epidemiology through a workshop and reach consensual understanding on the terms used to represent research quality in this project.

### 1 Research landscapes

The first phase of the project will describe the characteristics of the reported research project such as team members and the journal that publishes the article. The following lists the research characteristics of interest (See Supplementary Table 1)

a. Institution and qualification of the corresponding author/s
b. Numbers of authors, institutions and specialties
c. Numbers of oversea collaborating authors and institutions
d. Numbers of study site
e. Journal type: local vs. regional vs. international, open access vs. traditional subscription-based, general vs. discipline-specific
f. Setting-healthcare facility (hospital, clinic, etc.) or community
g. Type of study-audit vs. research-secondary (reviews) or primary (diagnostic, prognostic, etiologic or interventional), clinical vs. non-clinical (laboratory, public health, health service, etc.)
h. Data collection designs
i. Years when the study conducted, completed and published
j. Health conditions studied or organ systems that are involved
k. Drugs, devices/tools, surgical, psychological, or health services

### 2 Research quality

In phase 2 of the study, the research quality will be assessed based on the following criteria in three domains: relevance, credibility and usefulness (Table 1). At the start of screening, we will implement a training session for all reviewers in which all reviewers will extract the same articles. This will help ensure uniformity in the terminology and domains used. We will also determine the inter-rater reliability agreement using Cohen’s kappa κ and intra-class correlation (ICC).The kappa κ is a measure of agreement between different observers beyond chance agreement.^27^ κ statistic will be computed separately for each domain’s item (0 or 1). The ICC will be used to assess the domains’ subtotal (3, 4 and 3) and the grand total score of the tool (Table 1).

**Table 1.**
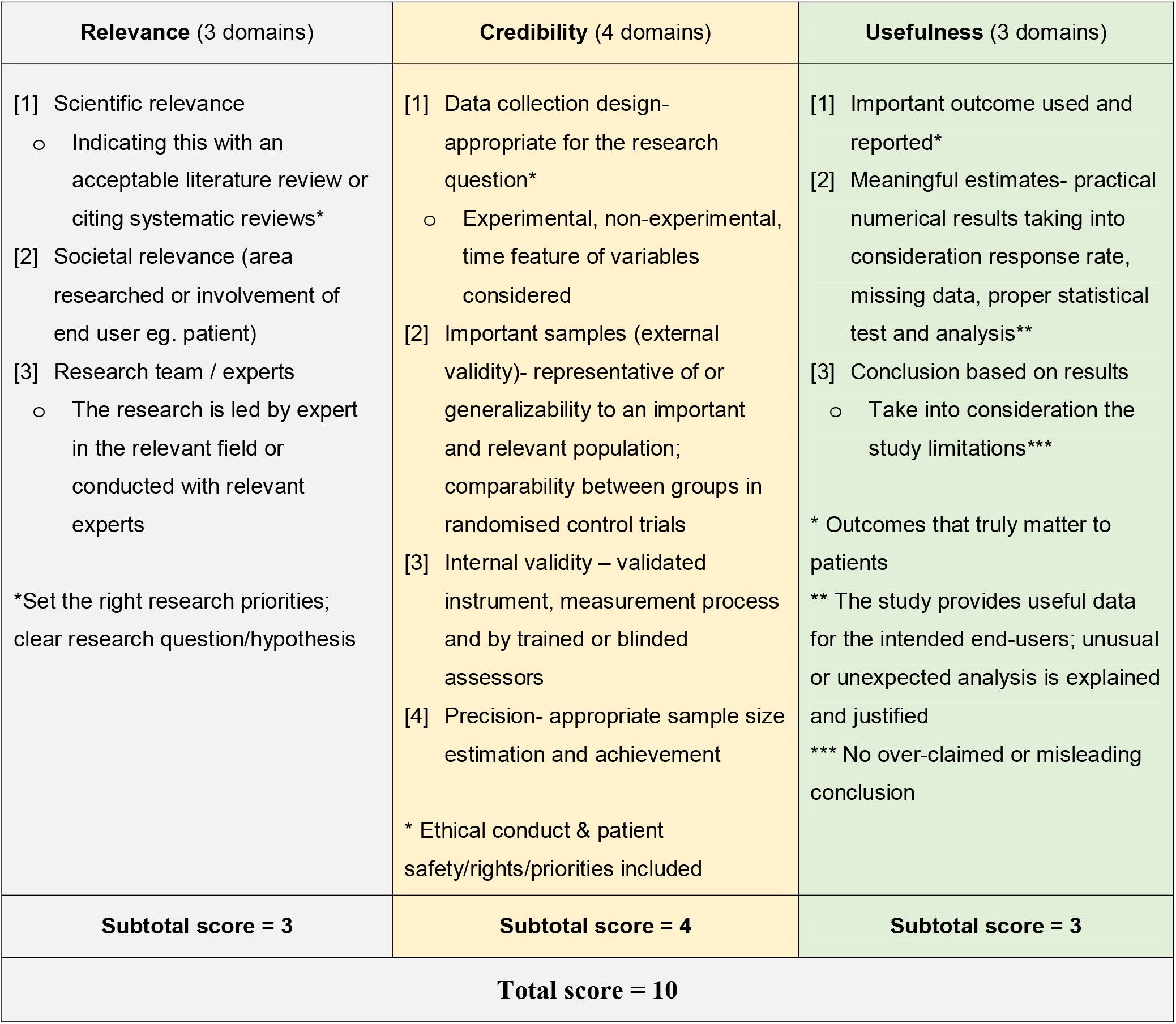
Research quality domains used in the screening tool

The Kappa result be interpreted as follows: values ≤ 0 as indicating no agreement and 0.01-0.20 as none to slight, 0.21-0.40 as fair, 0.4 - 0.60 as moderate, 0.61-0.80 as substantial, and 0.81-1.00 as almost perfect agreement.^28^ For the ICC, values < 0.40 is poor, 0 .40 - 0.59 is fair, 0.60 - 0.74 good, and 0.75 - 1.0 is excellent. We specify an *a priori* level of κ > 0.60 and ICC > 0.60 must be achieved before the second phase of this study begins. Retraining and reassessment of the reviewers on different articles will be conducted until the inter-rater agreement reach the desirable levels. The expected lower bound of a 95 % confidence limit for κ is no less than 0.60, with an assumed same marginal prevalence of zero score of 30%. Using alpha and beta error rates of 0.05 and 0.2, respectively, a pair of two reviewers will rate 20 papers each,^28,29^ with five pairs of reviewers and 100 samples for the subtotal and total ICC estimation.^30^

#### 2.1 Relevance

The relevance of a research will be assessed from three perspectives: scientific relevance, the composition of the research team and societal relevance. A research is being scientifically relevant if it addresses a true and real scientific problem and provides the needed knowledge to understand an existing phenomena. Scientific relevance also denotes that the research sets out on justified scientific foundation and informed of existing evidence. Thus, a scientifically relevant research is usually a globally relevant research due to its highly generalizable topic and subjects of research.

Societal relevance refers to the research that addresses a true and real problem in the society. This relevancy may exist at a smaller and wider population such as it may relevant for all the human population in the world or it may be relevant to a particular group of condition or disease in a unique population. These two domains of scientific and societal relevance relate to having a novelty in the research.

The last domain in the relevance category is about the research team of comprising investigators and experts of relevant professional qualifications. This may include patients and public people in certain research area when opinion of the end-users are considered important such as intervention or experience of the patients or family members.

#### 2.2 Credibility

This category is further assessed after it is judged that the research is relevant. Four essential features that are considered the very minimums in a research for it to be credible and its results to inform or contribute to practice change are data collection design, precision, important sample (external validity) and internal validity.

The design of the data collection of a research is to be appropriate to its objective or research question. The approach used in the data collection depends on whether it is a causal or non-causal research, and then experimental or non-experimental conduct of the research would provide better data. The time feature or characteristic of the variables involved in the research should be collected in their intended phases or stages such as a risk factor in the asymptomatic phase, or symptoms or biomarkers in the latent period.

Sampling and samples are the next important credibility domain. The sample of the participants is to be right group of the population for the research. They represent the important population to which the results could be generalised to later. However, in causal or experimental research, comparability between groups in the research take precedence over representativeness because confounding or prognostic factors between groups results in valid outcomes as of the exposure.

Quantitative research is essentially about measurement, measuring tools and process. The measurement of the variables is to be done by validated tools, through a standardised process, and if necessary by trained and blinded assessors. Any query or suspicion on the methods of measurement in the research will cause internal non-validity.

A credible research provides an appropriate and rational sample size estimation. This bases on the research question and its primary objective, and a similar earlier research. Adequate sample size is required for sufficient precision in a research. The achievement or non-achievement of the desired sample size should be reported or justified and discussed, respectively.

#### 2.3 Usefulness

The research that is credible worth its results a good attention. Usefulness of the research results consists of it being important outcomes, providing meaningful estimates and fair conclusion as supported by the research designs.

Important outcomes are that of high priority and concern to the end-users. These generally refer to the hard outcomes or strong correlates or intermediate markers of the hard outcomes to the exposure in the research. Examples of important outcomes include the diagnoses of the conditions, and the examples of the surrogates are blood or serum markers.

Results of a research are meaningful when they are easily understood in the context of clinical practice or daily life of patients. The meaningful estimates are usually the direct results or the research such as the actual numbers of occurrence, incidence and prevalence rates, and risk ratios. Transformed estimates such as standardised or log will need translation and interpretation.

Lastly, conclusion of the research bears the second testimony to that of the readers’ own judgement of the research. As the final interpretation and remarks by the authors and investigators of the research, it is important to put the results of the research as an evidence in the right context and applicability taken into consideration of the constraint in the research designs and limitations encountered along the whole research process.

## Data analysis

Every eligible article and study will be assessed according to two main areas – characteristics and quality of research. Data will be checked for any missing data and errors. The data will be reported descriptively, with frequency and percentage for categorical data while mean and standard deviation (median and interquartile range) for normally distributed (and not normally distributed) continuous data. Time series plot will be conducted to investigate the trends and patterns of the research characteristics, health conditions studied and quality of research over the years. Geographic information system (GIS) may also be plotted to evaluate the locations and areas of research conducted. Longitudinal trends of certain research characteristics, health conditions or areas in different settings will be explored.

Associations between characteristics of the included research project and quality will be explored, and the independent effect of each of the determinants will be quantified in multiple linear regression analysis. Additionally, the research quality as a categorical outcome will be explored as tertiles. The highest tertile will be compared to the lowest tertile, and the determinants will be assessed in multiple logistic regression. Longitudinal trends of the research quality will be explored. A calculated 95% confidence interval and two-sided α of 0.05 will be used to test significance. Model checking will be conducted in order to get the best and parsimony final model as well as fulfilled statistical assumptions. Estimates will be obtained with PASW 25.0 (SPSS, Chicago, IL) and MLwiN version 3.02 (Centre for Multilevel Modelling, University of Bristol).

## Discussion

Results will be informative to all stakeholders of clinical and biomedical research in the country of the evolution of research conduct and performance from the past till now. Profiles of the research throughout the past decades may be studied according to socioeconomic, politic or policy changes of certain years. The longitudinal and prospective trends of the research profiles, research quality and the association between them could provide suggestions on improvement initiatives or an institutional role model that has been ‘successful’ to some extent could be discovered. Additionally, health conditions or areas in different settings, and whether they are over- or under-studied may help future prioritization of research initiatives and resources. Descriptive comparison between countries may also be possible if there are similar studies done in other countries. This provides meaningful benchmarking and insights into the effects of evolving historical events on clinical and biomedical research activities and quality in each country.

The research quality tool of this study may be a useful screening tool for all quantitative study designs except qualitative study, case reports, and systematic reviews. We hope it would be a useful tool for a quick critical appraisal of research quality. The sequence of Relevance-Credibility-Usefulness enable efficiency and empower the tool users in the critical appraisal process. The main limitation of this review would be the reporting quality of the research including zero reporting or null publication of any completed studies.^32^ In addition, a relatively large number of graduate and postgraduate students’ research projects that were published as thesis and not in journals ^33^ will not be searchable through the search strategies used in this review project. Reporting quality is not assessed with the research quality tool that is created for this project because there are already specific guides and checklists for this purpose. The quality and comprehensiveness of the research reporting may be less worse than the research quality in terms of methodology but may affect its assessment.^34^ The 10 items within the three domains of the research quality screening tools are believed to be the fundamental minimums of most clinical and biomedical research that would be available in most published articles. Contacting the corresponding authors either through email or telephone would recover missing information in the included articles.

## Data Availability

Data may be shared upon completion of the review to any requesting party.

## Data Availability

Data may be shared upon completion of the review to any requesting party.

## Acknowledgments

We would like to thank the following colleague who have given helpful suggestion during the protocol development stage: Dr. Alfi Yasmina from the Faculty of Medicine, Lambung Mangkurat University, Banjarbaru, Kalimantan, Indonesia; Dr. Tan Kit-Aun from the Department of Psychiatry, Faculty of Medicine and Health Science, Universiti Putra Malaysia; Dr. Sanjiv Rampal Lekhraj from the Department of Orthopaedic, Faculty of Medicine and Health Science, Universiti Putra Malaysia; Dr. Dhashani Sivaratnam from the Unit of Ophthalmology, Department of Surgery, Faculty of Medicine and Health Science, Universiti Putra Malaysia; and Dr. Maliza Mawardi, Associate Professor Dr. Cheong Ai Theng, Associate Professor Dr. Lee Ping Yein and Associate Professor Dr. Ching Siew Mooi from the Department of Family Medicine, Faculty of Medicine and Health Science, Universiti Putra Malaysia.

